# Quantifying association of early proteinuria and eGFR changes with long-term kidney failure hazard in C3G and IC-MPGN

**DOI:** 10.1101/2024.02.03.24301605

**Authors:** Sherry Masoud, Katie Wong, David Pitcher, Lewis Downward, Clare Proudfoot, Nicholas J.A. Webb, RaDaR Consortium, Edwin K.S. Wong, Daniel P. Gale

**Author notes:** Multiple affiliations (RaDaR Consortium). Correspondence to: Professor Daniel P. Gale, UCL Department of Renal Medicine, Royal Free Hospital, Rowland Hill Street, London, NW3 2PF,.

## Abstract

**Background:** C3 glomerulopathy (C3G) and immune-complex membranoproliferative glomerulonephritis (IC-MPGN) are rare disorders that frequently result in kidney failure over the long-term. At present, there are no disease-specific treatments approved for these disorders, although there is much interest in the therapeutic potential of complement inhibition. However, the limited duration and necessarily small size of controlled trials means there is a need to quantify how well short-term changes in eGFR and proteinuria predict the clinically important outcome of kidney failure. We aimed to address this using longitudinal data from the UK National Registry of Rare Kidney Diseases (RaDaR).

**Methods:** RaDaR involves both retrospective and prospective data collection with linkage to hospital laboratories via automated feeds. 667 patients were included. Analyses of kidney survival were conducted using Kaplan–Meier and Cox regression. eGFR slope was estimated using linear mixed models.

**Results:** Over a median of 10.1 (IQR 6.9-14.3) years follow-up, 253/667 (38%) reached kidney failure. There was no difference in progression to kidney failure between C3G, IC-MPGN and Primary MPGN Not Otherwise Specified subgroups (p=0.75). Baseline urine protein creatinine ratio (UPCR), although high, was not associated with kidney failure risk. 2-year eGFR slope had a modest effect on kidney failure risk. In contrast, both 20-50% and 0.44g/g (50mg/mmol) reductions in time-averaged UPCR at 12 months were strongly associated with lower kidney failure risk (p≤0.002). Most notably, those with a UPCR <0.88g/g (<100mg/mmol) at 12 months had a substantially lower risk of kidney failure (HR 0.15 (95%CI 0.05-0.41).

**Conclusions:** We quantified the relationships between early changes in both eGFR and proteinuria with long-term kidney survival. We demonstrate that proteinuria a short time after diagnosis is a strong predictor of long-term outcome and that a UPCR <0.88g/g (<100mg/mmol) at 1 year is associated with a substantially lower kidney failure risk.

## Introduction

Primary membranoproliferative glomerulonephritis (MPGN) can be divided into C3 glomerulopathy (C3G) and immune-complex membranoproliferative glomerulonephritis (IC-MPGN). These are rare kidney disorders in which there is glomerular inflammation resulting in increased mesangial matrix and cellularity, capillary wall thickening with deposition of immunoglobulins (in IC-MPGN) and/or complement C3 (seen in both C3G and IC-MPGN). C3G is further subdivided into C3 glomerulonephritis and dense deposit disease (DDD) based on electron micrographic appearances with DDD characterized by dense transformation of the glomerular basement membrane.^1^ Presentation is usually with proteinuria and/or other features of kidney disease such as hematuria, hypertension, or renal impairment. Both conditions are rare with a combined incidence of 3-5 per million population.^2,3^ Although the biopsy features of C3G and IC-MPGN can also be seen in a range of disorders in which there is sustained activation of the immune system (such as persistent infection or autoimmune disease) the diagnosis of primary MPGN in the UK is reserved for those cases in which an underlying cause of immune activation is not identified. Although in most cases, the cause of C3G and IC-MPGN is not known, abnormal activation of the complement alternative pathway is frequently present in both disease categories. This can be attributed to the development of autoantibodies, the most common being C3 Nephritic Factor or, less often, can result from Mendelian^4–10^ or non-Mendelian rare or common genetic variants^11–15^ affecting innate or adaptive immunity; with comparable prevalence of variants and autoantibodies reported in both disorders.^12,16,17^ Together with the presence of C3 deposited in the kidneys in almost all cases and the frequent serological evidence of C3 consumption,^11,12,16^ these data have provided a compelling rationale for therapeutic targeting of the complement system in these disorders.

While the clinical presentation and diagnosis of these disorders are now well-established, long-term outcomes and prognostic features are less well understood, with the literature dominated by small, single center series, or studies with limited follow-up primarily focusing on baseline predictors of disease progression and prone to ascertainment bias. Nonetheless, certain prognostic markers have consistently been shown to be associated with kidney failure (KF) in both adult and pediatric IC-MPGN and/or C3G cohorts. These include baseline eGFR and hypoalbuminemia, along with renal biopsy findings of interstitial fibrosis and tubular atrophy, crescents, and segmental sclerosis.^12,18–24^ Literature regarding baseline proteinuria is more conflicting: in a study of 156 patients with C3G or IC-MPGN, baseline proteinuria >2g per day was independently associated with the composite outcome of doubling of serum creatinine or KF.^20^ However, in a cohort of 111 C3G patients from the US and 164 from France the association of baseline proteinuria with KF was non-significant in the multivariable model.^18,25^ Finally, the GLOSEN investigators demonstrated a ≥50% decrease in proteinuria over course of follow-up or within 6 and 12 months to be associated with a lower risk of KF.^26^

While case series and small observational studies have suggested a potential benefit of corticosteroids and mycophenolate, it is not known why some patients seem to respond and others do not, and long-term outcomes remain poor. The community therefore awaits with great expectation the results of several complement inhibition randomized trials. However, interpreting the potential clinical impact of an intervention for rare kidney diseases based on evidence of efficacy in a short (i.e. 0.5-2 year) clinical trial is often hampered by lack of direct data demonstrating efficacy in reducing the key clinically relevant outcome of KF. Therefore, data to inform appraisal of the likely clinical impact of early surrogate endpoints (such as proteinuria and short-term changes in eGFR) on long-term outcomes (such as KF) are needed by the nephrology community.^27–29^ Indeed recently there has been much interest in the extent to which these endpoints, amenable to study in relative short duration trials with limited numbers of participants, can serve as reliable surrogates for hard kidney outcomes and thus inform regulatory decisions and healthcare resource utilization. Both observational data and meta-analyses of controlled trial treatment effects have supported the use of eGFR slope,^30–32^ and proteinuria in the context of CKD,^33^ and IgA nephropathy.^34,35^ Subsequently, IgA therapies that demonstrate a short-term reduction in proteinuria in clinical trials can now apply for accelerated approval by the US Federal Drug Administration with full approval granted via a post marketing confirmatory trial using eGFR slope.

To address this unmet need in primary MPGN (C3G and IC-MPGN), here we use longitudinal data from 667 patients enrolled in the UK National Registry of Rare Kidney Diseases (RaDaR) to analyze and quantify the relationships between the clinical parameters of proteinuria and eGFR early in the disease course with the clinically important outcome of KF over the long-term. Therefore, this paper addresses and quantifies the first tenet for surrogate endpoints put forth by Prentice (which is that a surrogate endpoint should have a strong association with a true endpoint).^36^ The subsequent tenet, namely that a treatment effect on the surrogate must capture the treatment effect on the clinical end point is beyond the scope of this study and in general best achieved through meta-analysis of controlled trials.^32,37^ Additionally, while medication data enrichment for the primary MPGN cohort within RaDaR is ongoing, current data limitations preclude robust analyses of therapies patients have been exposed to historically.

## Methods

### Data source

RaDaR recruits patients from 108 NHS sites with both retrospective and prospective data collection through linkage with hospital laboratories for routine blood and urine test results via the UK Renal Data Collaboration, and with the UK Renal Registry (UKRR) for validated data on initiation of kidney replacement therapy (KRT), which includes data provided by NHS Blood and Transplant (NHSBT). Patients provide written informed consent at the time of recruitment. Details of recruitment characteristics and analyses of potential biases have been reported previously.^38^

Inclusion criteria for RaDaR are: child or adult with histological finding of: MPGN Type I; DDD (morphological pattern may or may not be MPGN); Other pattern of MPGN; C3 glomerulonephritis (characterized by C3 deposits in the absence of immunoglobulin with electron dense deposits (morphological pattern may or may not be MPGN); Unclassified GN with C3 dominant capillary wall immune deposits.

### Study population

Data from all prevalent patients recruited to RaDaR with one of the above conditions and diagnosed between January 2000 (from when proteinuria reporting to RaDaR was established) and December 2021 were extracted on 1st April 2024. Participants with an eGFR <15ml/min/1.73m^2^ or receiving kidney replacement therapy (KRT) at the time of diagnosis were excluded as were those with a known paraprotein (Figure 1). A subset of patients could be reliably classified as either C3G (n=153) or IC-MPGN (n=152), due to central histopathology review carried out for a previous pediatric observational study ^19^ or simultaneous review of available clinical and histopathology records within RaDaR (Figure 1). These 2 subgroups were compared to the remainder of participants with primary MPGN not otherwise specified (primary MPGN-NOS), with disease category specific analyses presented in supplementary materials.

**Figure 1.**
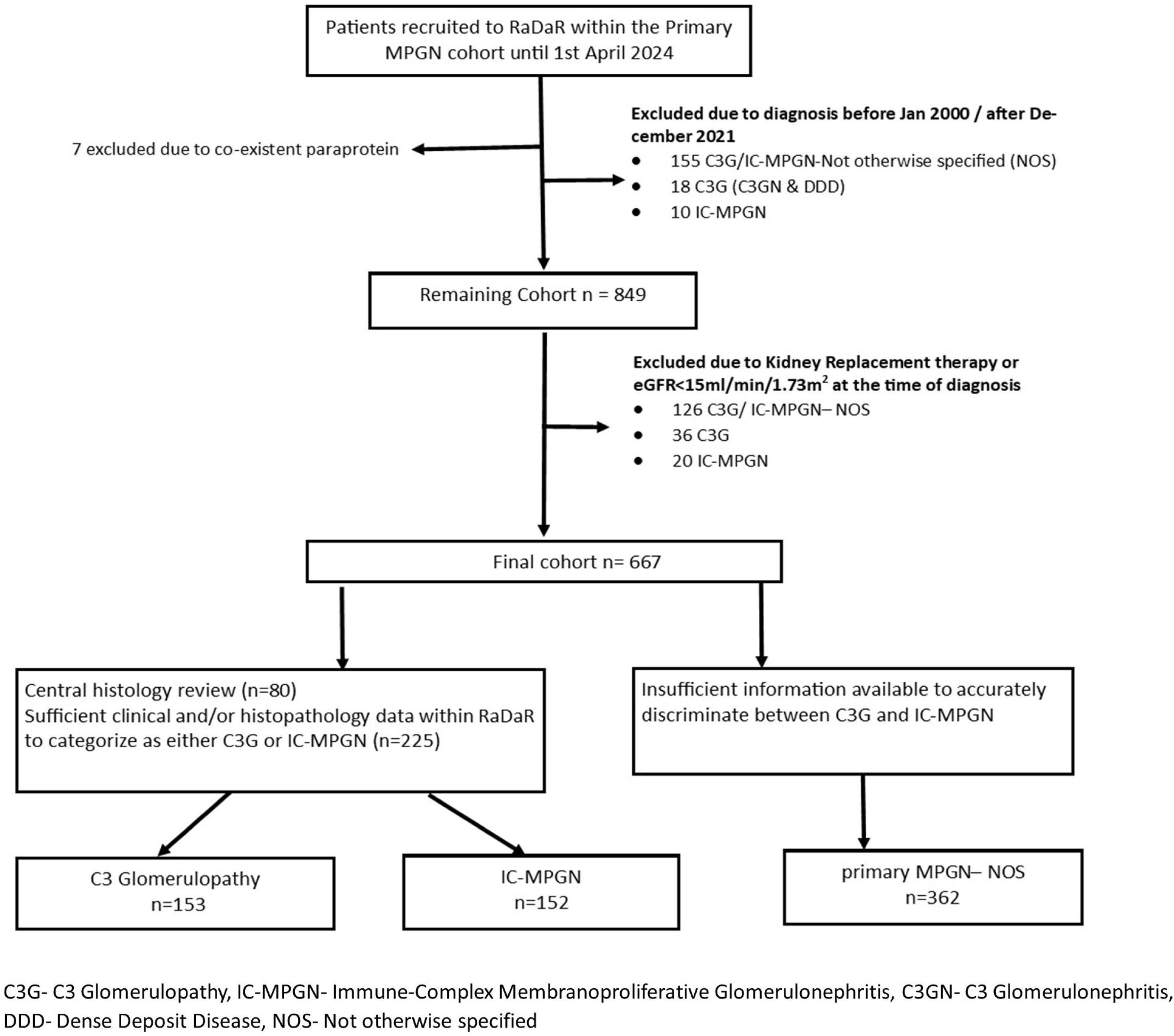
Study Flow Diagram

### Variable and outcome definitions

Baseline or diagnosis date was defined by kidney biopsy date or in the absence of this, date of diagnosis recorded in RaDaR. Time of diagnosis window was defined as +/-3-months from diagnosis date. eGFR was calculated from creatinine results using CKD-EPI (2009) without race adjustment, or Schwartz equation for those ≤16 years.^39,40^ Kidney failure (KF) was defined as dependance on KRT or eGFR ≤15mL/min/1.73m^2^ maintained for at least 4 weeks.^41^ Follow-up time was defined as time between date of diagnosis and last available test result, or whichever occurs first, KF or death from any cause.

### Statistical Analyses

Categorical data were reported as frequencies (%) and medians (interquartile range) for non-normally distributed continuous data. Kaplan-Meier analyses were used to compare time to KF for C3G, IC-MPGN and primary MPGN–NOS subgroups. Univariable Cox modelling was used to identify predictors of KF for the full cohort. Variables specified a priori included: age, sex, chronic kidney disease (CKD) stage, complement C3 and 4, random urine protein creatinine ratio (UPCR) at diagnosis and at 12months, immunosuppression within 1^st^ year of diagnosis, and primary MPGN sub-group (C3G, IC-MPGN or primary MPGN-NOS). Variables that achieved a significance threshold of p<0.20 were included in the final multivariable model. A two-sided p-value of 0.05 was considered significant. Cox regression was also used to investigate changes in UPCR over time, comparing diagnosis, 6-month, and 12-month timepoints with time to KF. Both percentage change in UPCR and an absolute reduction of 0.44g/g (50mg/mmol) in time averaged UPCR (TA-UPCR) were examined, with adjustment for sex, age, UPCR and eGFR at diagnosis. A reduction of 0.44g/g (50mg/mmol) was selected to examine the lowest prognostically meaningful change in UPCR. Progression to KF comparing those achieving a UPCR above or below a range of thresholds (from 0.44g/g to 4g/g) at 12months was also assessed.

Annualized rate of eGFR loss (eGFR slope) was calculated over full duration of follow-up, comparing C3G, IC-MPGN and primary MPGN-NOS sub-cohorts, and for the first two years following diagnosis. A linear mixed model with a minimum of four observations, random intercept and slope was used to estimate each patient’s eGFR slope. The association of KF with eGFR slope over 2 years and with percentage change in eGFR at 2 years (sustained over a minimum of 90 days) was also investigated, adjusting for age, sex, and eGFR at diagnosis. Finally, the impact of eGFR variability on KF, as measured using the coefficient of variation (CV) and average real variability (ARV), was evaluated, and adjusted for the same covariates.

Analyses were performed using SAS v9.4 and R v4.3.3.

### Ethics

This report adheres to the Strengthening the Reporting of Observation Studies in Epidemiology (STROBE) statement. RaDaR has ethical approval as a research registry provided by NHS South-West Central Bristol Research ethics committee (14/SW/1088) and by the RaDaR and UKRR operational committees.

## Results

### Demographics and baseline characteristics

667 patients were included, 362 (54%) with primary MPGN-NOS, 153 (23%) with definitive C3G and 152 (23%) with definitive IC-MPGN (Figure 1). Both definitive C3G and IC-MPGN subgroups had a higher proportion of pediatric participants (Table 1). Participant characteristics further stratified by C3G subtype (C3GN or DDD) can be found in Supplemental Table 1. The median age of the overall cohort at diagnosis was 35 years (IQR 15-56) with 207/667 (31%) diagnosed at <18 years old. Approximately half of the participants were female (46%), and most were white (89%); which was similar across sub-groups. Kidney function at diagnosis was relatively preserved with a median eGFR of 67 mL/min/1.73 m^2^ (IQR 38-103) and significant proteinuria at baseline UPCR 3.8g/g (IQR 1.3-7.2). Median C3 levels were lowest in the definitive C3G subgroup (0.23g/L (0.12-0.43), with comparable C4 levels across subgroups. At least one medication entry was available for 487/667 participants within the 1^st^ year of diagnosis, of those with data 138/487 (28%) received at least one immunosuppressant and 128 (26%) received corticosteroids alone or as combination therapy. 125/487 (26%) were recorded as receiving a renal angiotensin system (RAS) inhibitor within 1 year of diagnosis, however this could reflect incomplete medication data collection.

**Table 1.** Baseline characteristics and clinical outcomes.

|  | C3 Glomerulopathy |  | IC-MPGN |  | Primary MPGN-Not Otherwise Specified |  | Overall Cohort |  |
| --- | --- | --- | --- | --- | --- | --- | --- | --- |
|  | N=153<br>(C3GN =99<br>DDD =54) | (%)* | N=152 | (%)* | N=362 | (%)* | N=667 | (%)* |
| Age at diagnosis (years) |  |  |  |  |  |  |  |  |
| Median (IQR) | 17 (11 - 40) |  | 24 (10 - 53) |  | 43 (23 – 61) |  | 35 (15-56) |  |
| Pediatric (<18 years) | 79 | (52) | 70 | (46) | 58 | (16) | 207 | (31) |
| Sex |  |  |  |  |  |  |  |  |
| Female | 66 | (43) | 73 | (48) | 165 | (46) | 304 | (46) |
| Ethnicity |  |  |  |  |  |  |  |  |
| White | 113 | (84) | 125 | (89) | 275 | (90) | 513 | (89) |
| Median follow up duration |  |  |  |  |  |  |  |  |
| Median (IQR), years | 9.8 (7.5 - 15.8) |  | 11.5 (8.0 – 14.7) |  | 9.5 (6.5 – 14.0) |  | 10.1 (6.9-14.3) |  |
| UPCR (Median IQR, g/g) |  |  |  |  |  |  |  |  |
| Diagnosis | 3.6 (1.0 - 6.7) |  | 4.3 (1.6 - 7.4) |  | 2.8 (0.8 - 7.0) |  | 3.8 (1.2 - 7.3) |  |
| 6-month | 1.3 (0.6 - 3.7) |  | 0.9 (0.2 – 2.6) |  | 1.6 (0.3 – 3.8) |  | 1.3 (0.4 - 3.5) |  |
| 12-month | 1.2 (0.3 - 3.8) |  | 0.5 (0.2 – 2.9) |  | 0.7 (0.2 – 2.8) |  | 0.8 (0.2 - 3.1) |  |
| eGFR at diagnosis |  |  |  |  |  |  |  |  |
| Median (IQR), mL/min/1.73 m<sup>2</sup> | 82 (46 - 106) |  | 68 (42 - 118) |  | 55 (33 - 90) |  | 67 (38 -103) |  |
| Serum albumin at diagnosis |  |  |  |  |  |  |  |  |
| Mean (SD), g/dL | 3.1 (0.9) |  | 2.8 (0.9) |  | 3.1 (0.9) |  | 3.0 (0.9) |  |
| Complement C3 levels at diagnosis |  |  |  |  |  |  |  |  |
| Median (IQR), g/L | 0.23 (0.12 - 0.43) |  | 0.64 (0.17 - 0.97) |  | 0.61 (0.18 – 0.75) |  | 0.41 (0.16-.073) |  |
| Complement C4 levels at diagnosis |  |  |  |  |  |  |  |  |
| Median (IQR), g/L | 0.19 (0.14 - 0.28) |  | 0.14 (0.08 - 0.21) |  | 0.18 (0.12 – 0.25) |  | 0.19 (0.11-0.26) |  |
| Kidney failure event |  |  |  |  |  |  |  |  |
| Yes | 60 | (39) | 56 | (37) | 137 | (38) | 253 | (38) |
| Immunosuppression within 1 year of diagnosis |  |  |  |  |  |  |  |  |
| Yes | 47 | (40) | 47 | (42) | 45 | (17) | 139 | (29) |
| RAS inhibitor within 1 year of diagnosis |  |  |  |  |  |  |  |  |
| Yes | 46 | (39) | 40 | (36) | 39 | (15) | 125 | (26) |

\*Percentages are proportions of those with data available. C3GN-C3 Glomerulonephritis, DDD- Dense Deposit Disease, IC-MPGN- Immune-Complex Membranoproliferative Glomerulonephritis, Primary MPGN- Primary Membranoproliferative Glomerulonephritis, IQR- interquartile range, eGFR – estimated glomerular filtration rate, UPCR- urine protein creatinine ratio, RAS- renal angiotensin system.

### Kidney Replacement Therapy

Of the 223 who started KRT over the course of follow-up, 36/223 (16%) were diagnosed at <18 years old. Most began KRT on maintenance hemodialysis 15/36 (42%), followed by 13/36 (36%) on peritoneal dialyses and 7/36 (19%) who received a pre-emptive kidney transplant. For those diagnosed in adulthood, the first modality was hemodialysis for 106/187 (57%), peritoneal dialysis for 55/187 (29%) and pre-emptive transplant for 26/187 (14%). Over the follow-up period 120/187 (64%) adults and 30/36 (83%) children underwent at least one kidney transplant. When also including those with CKD5 at diagnosis, the total increases to 285 participants, with a median time to graft failure of 6.5 years (95%CI 4.7-8.1), and figures of 6.5 years (2.5-8.3) for C3G and 6.0 years (4.7-11.9) for IC-MPGN (Supplemental Figure 1). Median time to graft failure for all subsequent transplants was 3.3 years (95%CI 0.9 - Non calculable).

### Risk Factors for Progression to Kidney Failure

Over a median follow-up of 10.1 years (IQR 6.9-14.3), 253/667 (38%) progressed to kidney failure (KF). Linear mixed models of eGFR slope over full duration of follow-up and Kaplan-Meier analyses demonstrated no statistically significant difference in progression to KF between definitive C3G, definitive IC-MPGN and primary MPGN-NOS sub-groups (Figure 2). All subsequent analyses were performed on the overall cohort, with disease specific analyses presented in supplemental materials.

**Figure 2.**
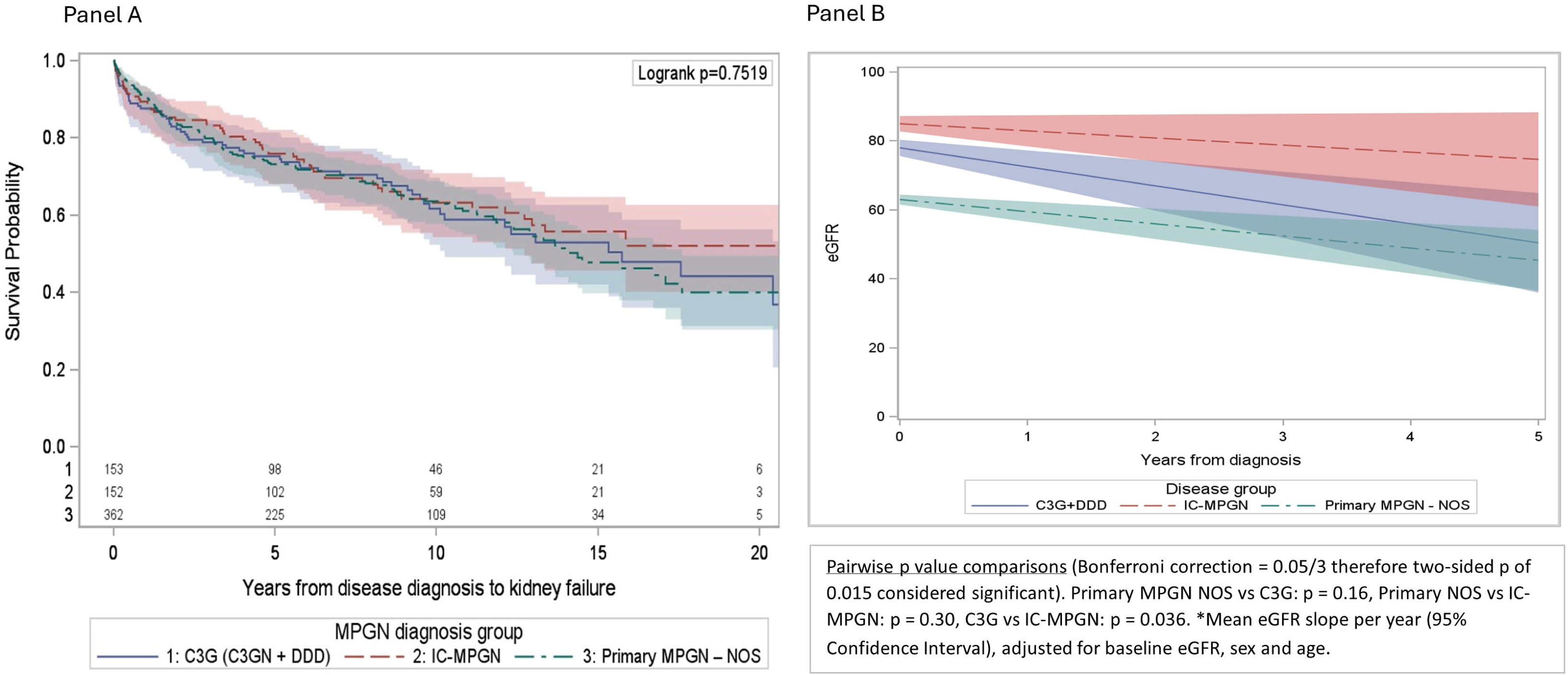
Kaplan-Meier of time to kidney failure by disease subgroup (Panel A). Adjusted eGFR slope over full duration of follow-up truncated at 5 years by disease subgroup (Panel B)

Predictors of progression to KF were investigated using univariate and multivariable models (Table 2). CKD stage at diagnosis and UPCR levels of 0.88-2.64g/g or >2.64g/g at 12 months were independent predictors of KF, whereas UPCR category at diagnosis, use of immunosuppression within 1 year, complement C3 and C4, age, sex, and disease subgroup were not. We repeated the analysis with a limited model which included UPCR and eGFR at diagnosis as continuous variables, age and sex, again UPCR at diagnosis was not an independent predictor of outcome (p>0.2).

**Table 2.**
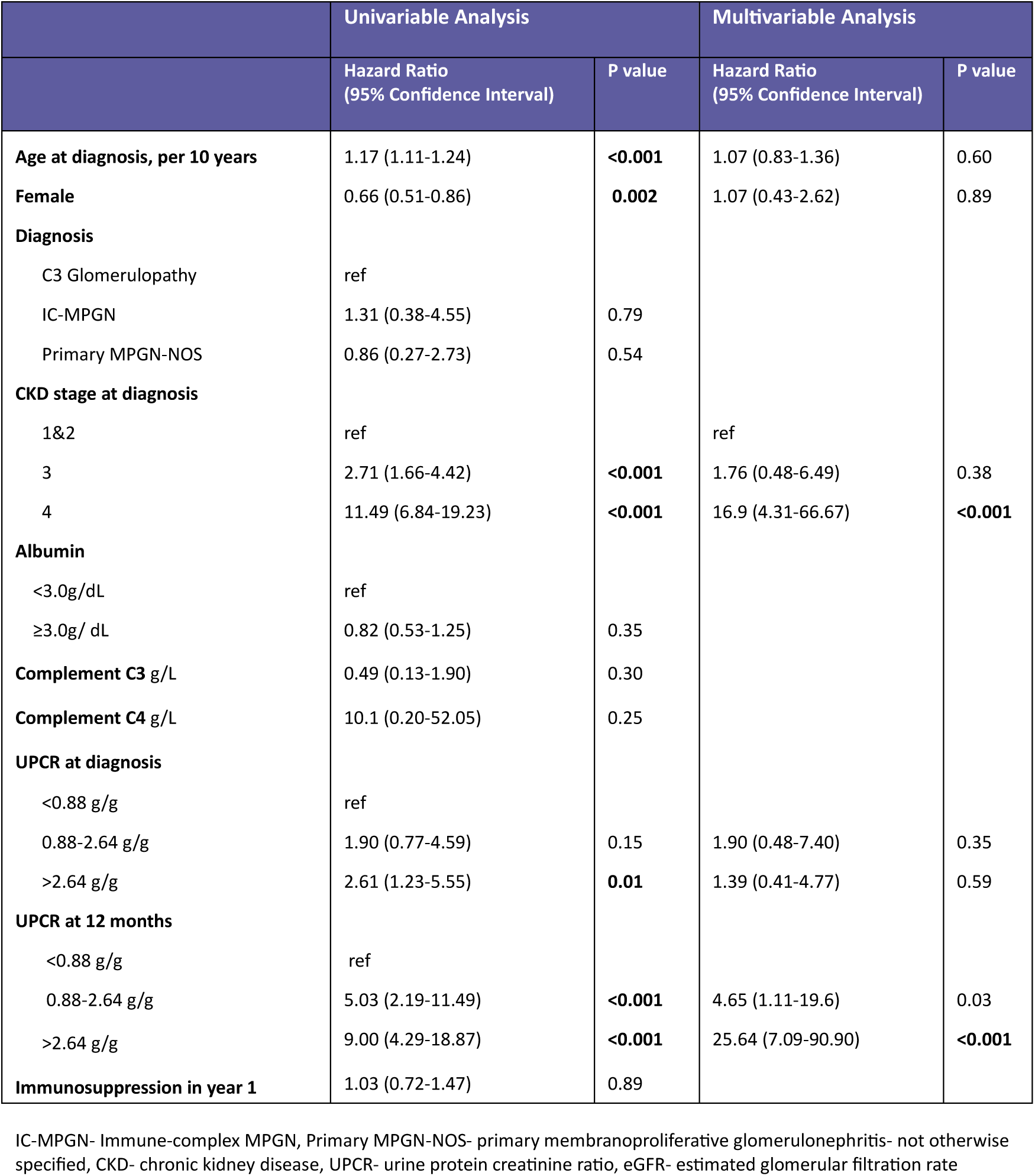
Univariable and multivariable cox model of kidney failure predictors.

Addressing whether changes in eGFR early in disease course can predict the later development of KF, we first demonstrate that annualized eGFR slope calculated over the first 2 years following diagnosis was strongly associated with KF (p<0.001) (Figure 3A. Supplemental Figure 2). However, an annual decline of 10ml/min/1.73m^2^ over the first 2 years was associated with only a modest increase in kidney failure hazard (HR 1.69 95%CI 1.49-1.92). This was robust to sensitivity analyses excluding those with an eGFR >60ml/min/1.73m^2^ at diagnosis, with subsequent point estimates only marginally higher (Figure 3B). Replicating this in a prevalent cohort (diagnosed >1 year prior to inclusion) resulted in higher point estimates, however confidence intervals overlapped with those of the initial incident or early disease cohort (Supplemental Figure 3). The more typical clinical trial endpoint of (sustained) percentage change in eGFR at 2 years, was also strongly associated with KF (Figure 3C), with the distribution of participants’ eGFR changes available in Supplemental Figure 4. eGFR variability as measured by both CV and ARV was not associated with kidney failure (p>0.30).

**Figure 3.**
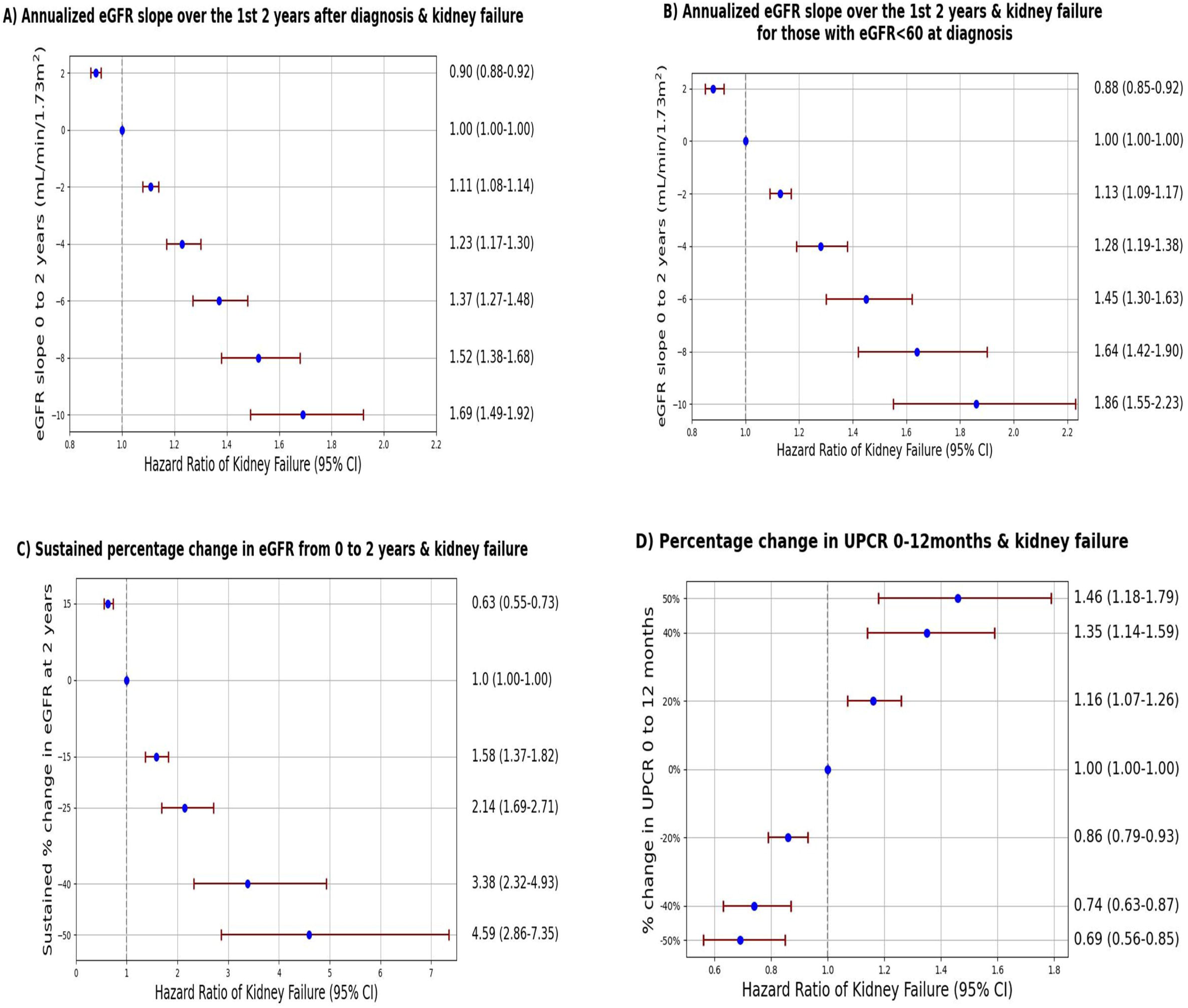
Forest plots of UPCR and eGFR changes within 2 years of diagnosis and hazard ratio of kidney failure

Next, we examined changes in UPCR across diagnosis, 6-month, 12-month timepoints as may be presented in a clinical trial, excluding those with a UPCR <0.44g/g (50mg/mmol) at diagnosis. As outlined, the objective was to quantify the KF hazard associated with increases and decreases in UPCR, regardless of why these may have occurred. The distribution of UPCR measurements in our cohort across different timepoints can be found in Supplemental Figure 5 with median UPCR of 3.8g/g (IQR 1.2-7.3) at diagnosis and 0.8g/g (IQR 0.2-3.1) at 12months. We examined absolute changes in TA-UPCR to account for variability between measurements, and a reduction as little as 0.44g/g was significantly associated with lower risk of KF at a consistent magnitude for all timepoints 0-6months, 0-12months and 6-12months (p ≤0.003, Table 3, Supplemental Table 2). Additionally, whilst a 50% reduction in UPCR at 6 months did not reach statistical significance - HR 0.88 (95%CI 0.72-1.07), a halving of UPCR from diagnosis to 12 months (HR 0.69 (0.56-0.85)) and 6 to 12months (HR 0.75 (0.65-0.86)) was strongly associated with a lower rate of KF (Table 3, Supplemental Table 2). Forest plots demonstrating how this risk varies for a range of UPCR changes (20-50%) from diagnosis to 12 months can be found in Figure 3D (Supplemental Figure 6), with the distribution of UPCR changes in our cohort presented in Supplemental Figure 7.

**Table 3.** UPCR thresholds and changes in UPCR in the 1^st^ 12 months following diagnosis and risk of kidney failure for the full cohort.

| UPCR thresholds at 12 months and Hazard Ratio (95% Confidence Interval) of Kidney Failure |  |  |  | 50% decline in UPCR and Hazard Ratio (95% Confidence Interval) of Kidney Failure * |  |  |  |  | 0.44g/g (50mg/mmol) decline in UPCR and Hazard Ratio (95% Confidence Interval) of Kidney Failure* |  |  |  |  |
| --- | --- | --- | --- | --- | --- | --- | --- | --- | --- | --- | --- | --- | --- |
| UPCR Threshold | Adjusted HR**<br>N=117 | Adjusted HR**<br>(sensitivity analysis<sup>1</sup>)<br>N=78 | P value<br>(both analyses) | Timepoint from | Timepoint to | N | Adjusted HR<sup>II</sup> | P value | Timepoint from | Timepoint to | N | Adjusted HR<sup>II</sup> | P value |
| <0.88g/g | 0.15 (0.05-0.41) | 0.05 (0.006-0.42) | <0.001 | Diagnosis | 6 months | 94 | 0.88 (0.72-1.07) | 0.21 | Diagnosis | 6 months | 99 | 0.90 (0.84-0.96) | 0.002 |
| <1.74g/g | 0.19 (0.09-0.44) | 0.17 (0.06-0.50) | <0.001 | Diagnosis | 1 year | 81 | 0.69 (0.56-0.85) | 0.002 | Diagnosis | 1 year | 83 | 0.89 (0.83-0.96) | 0.003 |
| <2.64g/g | 0.32(0.16-0.66) | 0.28 (0.12-0.68) | <0.001 | 6 months | 1 year | 75 | 0.75 (0.65-0.86) | <0.001 | 6 months | 1 year | 79 | 0.87 (0.78-0.96) | <0.001 |

UPCR- urine protein creatinine ratio, HR- hazard ratio. \*Excludes those with a UPCR <0.44g/g at diagnosis, \*\* Adjusted for age, sex and eGFR, <sup>1</sup>analysis excludes those with UPCR <0.88g/g at diagnosis (or low risk disease), <sup>II</sup>Adjusted for eGFR, age, sex and baseline UPCR

From both a clinical and trial perspective, treatment targets and to what extent reaching these targets diminishes KF risk can often be useful. Figure 4 shows time to KF according to UPCR category and Table 3 shows the KF hazard for those who do and do not achieve a specific threshold of UPCR at 12 months irrespective of starting UPCR. For example, a UPCR less than 0.88g/g (100mg/mmol) at 12 months was associated with an 85% lower rate of KF. To verify this finding was not driven by inclusion of low-risk participants whose UPCR started low and remained low, we performed a sensitivity analysis excluding all those with a UPCR <0.88g/g at diagnosis. All three calculated hazard ratios were lower than those of the original analyses (Table 3). Figure 5 provides a visual depiction of the hazard ratios of KF above and below a range of UPCR thresholds plotted on the x axis. Notably the most pronounced changes in risk occur up to a threshold of 1g/g and thereafter a relative attenuation in the hazard ratio is evident. In addition, the strikingly low risk of kidney failure associated with a UPCR < 0.88g/g at 12 months was also replicated in C3G only subgroup (HR 0.14 (0.05-0.52)) with insufficient events in IC-MPGN subgroup.

**Figure 4.**
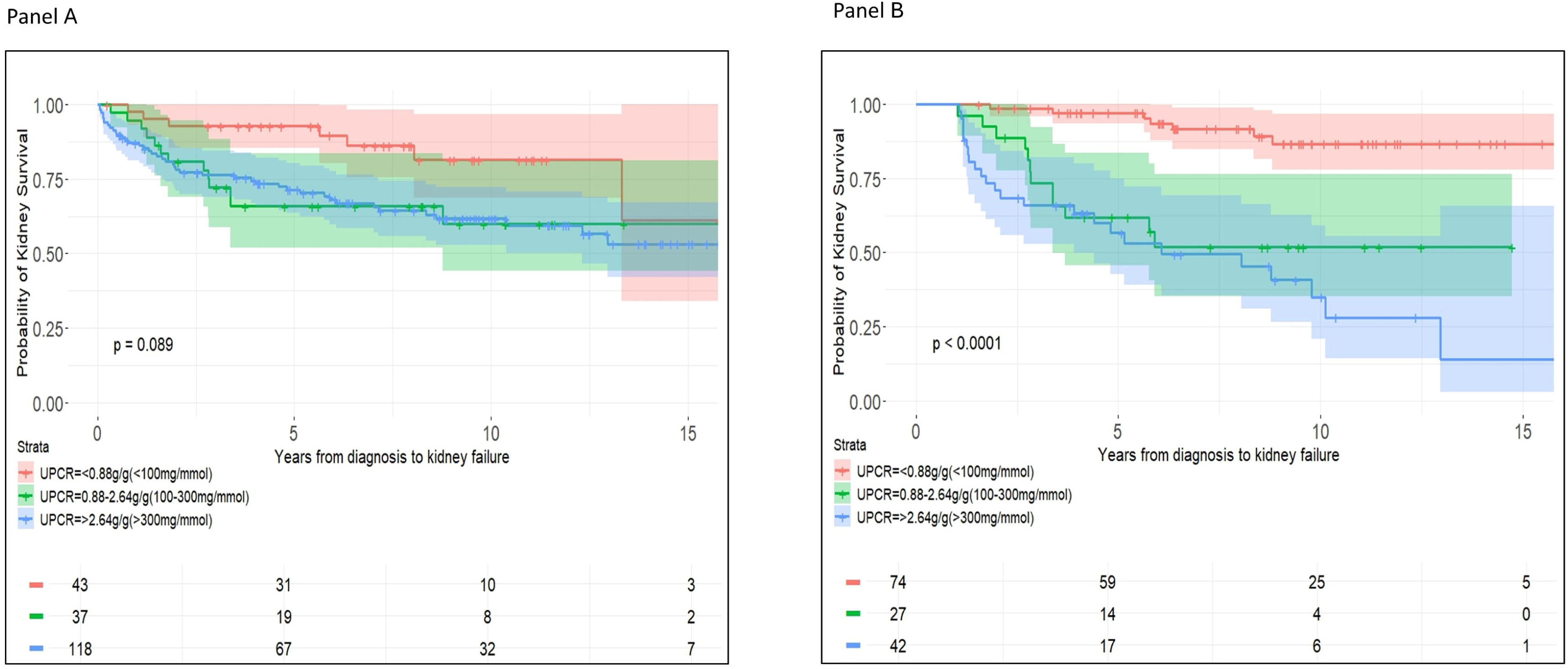
Kaplan-Meier of time to kidney failure according to UPCR category at diagnosis (Panel A) and UPCR category at 12 months (Panel B)

**Figure 5.**
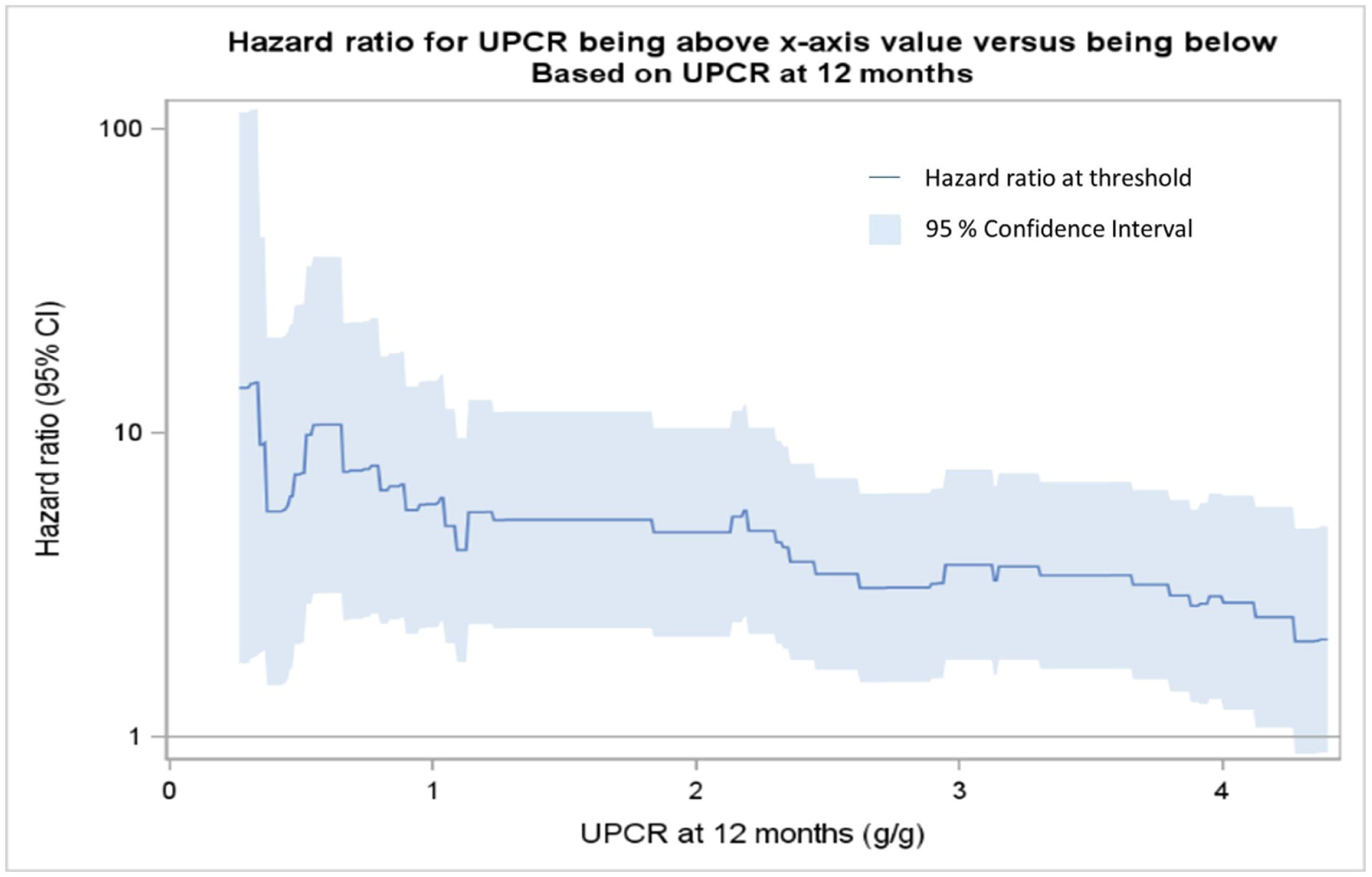
Adjusted hazard ratio of kidney failure on a log scale across a range of UPCR thresholds at 12 months

## Discussion

Herein we present long-term longitudinal data from a large cohort of 667 C3G or IC-MPGN patients within the UK National Registry of Rare Kidney Diseases (RaDaR). The findings provide insight into the natural history of these disorders, expanding on earlier, small-scale observational studies and providing quantitative estimates for the relationship of early surrogate endpoints on KF hazard. We present analyses of C3G and IC-MPGN both combined and separately for reference, given evidence of overlapping pathogenesis, specifically alternative complement pathway dysregulation and thus suitability for inclusion in targeted therapy trials.^11,12^ A particular strength of this study is the duration of follow-up, with a median 10.1 years (6.9-14.3), over which period 253/667 (38%) participants reached KF illustrating the significant unmet need for effective treatments in these disorders that cause KF at a young age.^42^ This also is notably compounded by the short time to first allograft loss (median 6.5 years (95%CI 4.7-8.1) compared with 84-87% five-year graft survival for (adult or pediatric) deceased donor recipients in the UK.^43,44^ Consistent with a number of studies,^12,45^ we show no significant difference in time to KF between C3G and IC-MPGN patients nor in mean eGFR slope over the first 5 years of follow-up. We demonstrate, as shown previously, that eGFR and proteinuria are strongly associated with long-term outcomes.^20,23^ However, our analysis also showed stronger relationships of these parameters at 6-24 months with long-term risk of KF, with the predictive value of proteinuria (and changes in proteinuria) particularly significant.

Addressing the utility of these endpoints in a disease specific context, we show that while eGFR slope early in disease course is strongly associated with KF, the magnitude of the effect is relatively modest, even over two years, compared to change in proteinuria over 1 year. This remains the case irrespective of whether baseline eGFR is above or below 60ml/min/1.73m^2^ as has also been shown using CKD data.^30^ This suggests that eGFR slope has more limited predictive power in the context of primary MPGN as compared to other kidney disorders, particularly early in the disease.

In contrast, whilst proteinuria at baseline was not associated with kidney failure as noted by others,^18,25^ TA-UPCR reduction by as little as 0.44g/g (50mg/mmol) was consistently associated with lower risk of KF, as was percentage change in UPCR. This complements previous reports from the GLOSEN registry, which showed a ≥50% reduction in proteinuria at 12 months was associated with a lower risk of KF (HR 0.83 95%CI (0.69–0.95)).^26^ We further extend this work and demonstrate smaller reductions in proteinuria i.e. 20-50% are also associated with reduction in the risk of KF. Additionally, achieving a threshold UPCR of <0.88g/g (<100mg/mmol) by 12 months was particularly strongly associated with lower rate of KF events (HR 0.15 (95%CI 0.05-0.41); p<0.001). Therefore, if proteinuria is shown in clinical trials to be reduced to similarly low levels by therapies that act by reducing disease activity, it is logical to infer that long-term KF hazard will be similarly reduced, supporting the use of this accessible endpoint in future trials as a surrogate for KF.

Our findings must be considered in the context of the limitations inherent in registry studies including incomplete data. The latter is mitigated through data linkages with UKRR and NHSBT which provide validated long-term KF endpoints for all UK patients as well as increasing prospective data collection via automated laboratory feeds from NHS hospitals. However, this remains a real-world dataset in which standard of care may impact the availability of eGFR and UPCR data at timepoints, as may patient or disease characteristics. It is most representative of the population and clinical practice patterns in the UK, which may be different in other settings. Additionally, medication data was limited, and the additive prognostic value of autoantibody or genetic variant status could not be assessed with this dataset. Finally, whilst beyond scope this study, further work to assess whether treatment effects on intermediate endpoints predict treatment effects on KF may enable upgrade of proteinuria from a “reasonably likely” to a “validated” endpoint as indicated in the Biomarkers, Endpoints, and other Tools Resource.^46^

In conclusion, using real-world longitudinal data from RaDaR, we provide quantitative descriptions of the relationships between early changes in both eGFR and proteinuria, and long-term renal outcomes in patients with C3G and IC-MPGN. Across a range of measures, we demonstrate that proteinuria a short time after diagnosis is a strong predictor of long-term outcomes and most notably that UPCR <0.88g/g (<100mg/mmol) at 1 year is associated with substantially lower risk of progressing to kidney failure.

## Acknowledgements

The authors thank all RaDaR participants, their family members, and the UK Renal Registry staff. RaDaR is funded by the UK Medical Research Council, Kidney Research UK, Kidney Care UK, and the Polycystic Kidney Disease Charity.

## Supplemental Material

Supplemental Table 1. Baseline characteristics and clinical outcomes of C3 Glomerulonephritis and Dense Deposit Disease

Supplemental Table 2. Urine protein creatinine ratio (UPCR) changes and hazard ratio of kidney failure definitive C3 and IC-MPGN subgroups

Supplemental Figure 1. Kaplan-Meier of time to 1^st^ kidney transplant failure comparing C3G, IC-MPGN and full cohort

Supplemental Figure 2. Forest plot of eGFR slope within 2 years of diagnosis and risk of kidney failure (KF) for definitive C3G subgroup (Panel A) and definitive IC-MPGN subgroup (Panel B)

Supplemental Figure 3. Forest plot of 2-year eGFR slope for full cohort of prevalent patients (minimum 1 year from diagnosis) with an eGFR <60ml/min/1.73m^2^ and risk of kidney failure

Supplemental Figure 4. Distribution of percentage change in eGFR between 0 and 2 years for full cohort

Supplemental Figure 5. Distribution of UPCR measurements for full cohort (Panel A) and on a log scale (Panel B)

Supplemental Figure 6. Forest plot of percentage in UPCR 0 to 12 months and unadjusted risk of kidney failure (KF) for definitive C3G subgroup (Panel A) and definitive IC-MPGN subgroup (Panel B)

Supplemental Figure 7. Distribution of percentage change in UPCR between 0 and 12 months for full cohort

## Authors Contributions

DPG conceived the study and acquired funding. SM, KW, LD, and DP curated data, performed formal analyses, accessed, and verified the data. DPG and SM wrote the initial draft. EW, CP, and NW assisted with analysis and reviewed and edited the manuscript. DPG and SM had final responsibility to submit for publication.

## Data availability

The RaDaR database is hosted by the UK Renal Registry and its metadata are available via https://ukkidney.org/rare-renal/homepage. Individual-level data are not available for export. Proposals to perform analyses using the data for academic, audit or commercial purposes can be made to the RaDaR Operations Group via https://ukkidney.org/rare-renal/homepage.

## References

1. Pickering MC, D’agati VD, Nester CM, et al. C3 glomerulopathy: consensus report. Kidney international. 2013;84(6):1079–1089.

2. Briganti EM, Dowling J, Finlay M, et al. The incidence of biopsy-proven glomerulonephritis in Australia. Nephrology Dialysis Transplantation. 2001;16(7):1364–1367.

3. Sethi S, Zand L, Leung N, et al. Membranoproliferative glomerulonephritis secondary to monoclonal gammopathy. Clinical journal of the American Society of Nephrology: CJASN. 2010;5(5):770.

4. Gale DP, De Jorge EG, Cook HT, et al. Identification of a mutation in complement factor H-related protein 5 in patients of Cypriot origin with glomerulonephritis. The Lancet. 2010;376(9743):794–801.

5. Malik TH, Lavin PJ, de Jorge EG, et al. A hybrid CFHR3-1 gene causes familial C3 glomerulopathy. Journal of the American Society of Nephrology: JASN. 2012;23(7):1155.

6. Chen Q, Wiesener M, Eberhardt HU, et al. Complement factor H–related hybrid protein deregulates complement in dense deposit disease. The Journal of clinical investigation. 2014;124(1):145–155.

7. Tortajada A, Yébenes H, Abarrategui-Garrido C, et al. C3 glomerulopathy–associated CFHR1 mutation alters FHR oligomerization and complement regulation. The Journal of clinical investigation. 2013;123(6):2434–2446.

8. Ault BH, Schmidt BZ, Fowler NL, et al. Human factor H deficiency. Journal of Biological Chemistry. 1997;272(40):25168–25175.

9. Martínez-Barricarte R, Heurich M, Valdes-Cañedo F, et al. Human C3 mutation reveals a mechanism of dense deposit disease pathogenesis and provides insights into complement activation and regulation. The Journal of clinical investigation. 2010;120(10):3702–3712.

10. Chauvet S, Roumenina LT, Bruneau S, et al. A familial C3GN secondary to defective C3 regulation by complement receptor 1 and complement factor H. Journal of the American Society of Nephrology: JASN. 2016;27(6):1665.

11. Servais A, Noël L-H, Roumenina LT, et al. Acquired and genetic complement abnormalities play a critical role in dense deposit disease and other C3 glomerulopathies. Kidney international. 2012;82(4):454–464.

12. Iatropoulos P, Noris M, Mele C, et al. Complement gene variants determine the risk of immunoglobulin-associated MPGN and C3 glomerulopathy and predict long-term renal outcome. Molecular immunology. 2016;71:131–142.

13. Bu F, Borsa NG, Jones MB, et al. High-throughput genetic testing for thrombotic microangiopathies and C3 glomerulopathies. Journal of the American Society of Nephrology: JASN. 2016;27(4):1245.

14. Levine AP, Chan MM, Sadeghi-Alavijeh O, et al. Large-scale whole-genome sequencing reveals the genetic architecture of primary membranoproliferative GN and C3 glomerulopathy. Journal of the American Society of Nephrology: JASN. 2020;31(2):365.

15. Afolabi H, Zhang BM, O’Shaughnessy M, Chertow GM, Lafayette R, Charu V. The Association of Class I and II Human Leukocyte Antigen Serotypes With End-Stage Kidney Disease Due to Membranoproliferative Glomerulonephritis and Dense Deposit Disease. American Journal of Kidney Diseases. 2024;83(1):79–89.

16. Iatropoulos P, Daina E, Curreri M, et al. Cluster analysis identifies distinct pathogenetic patterns in C3 glomerulopathies/immune complex–mediated membranoproliferative GN. Journal of the American Society of Nephrology. 2018;29(1):283–294.

17. Marinozzi MC, Roumenina LT, Chauvet S, et al. Anti-factor B and anti-C3b autoantibodies in C3 glomerulopathy and Ig-associated membranoproliferative GN. Journal of the American Society of Nephrology. 2017;28(5):1603–1613.

18. Bomback AS, Santoriello D, Avasare RS, et al. C3 glomerulonephritis and dense deposit disease share a similar disease course in a large United States cohort of patients with C3 glomerulopathy. Kidney international. 2018;93(4):977–985.

19. Wong EK, Marchbank KJ, Lomax-Browne H, et al. C3 glomerulopathy and related disorders in children: etiology-phenotype correlation and outcomes. Clinical Journal of the American Society of Nephrology. 2021;16(11):1639–1651.

20. Lomax-Browne HJ, Medjeral-Thomas NR, Barbour SJ, et al. Association of histologic parameters with outcome in C3 glomerulopathy and idiopathic immunoglobulin-associated membranoproliferative glomerulonephritis. Clinical Journal of the American Society of Nephrology. 2022;17(7):994–1007.

21. Caravaca-Fontan F, Trujillo H, Alonso M, et al. Validation of a histologic scoring index for C3 glomerulopathy. American Journal of Kidney Diseases. 2021;77(5):684–695. e1.

22. Kirpalani A, Jawa N, Smoyer WE, et al. Long-term outcomes of C3 glomerulopathy and immune-complex membranoproliferative glomerulonephritis in children. Kidney International Reports. 2020;5(12):2313–2324.

23. Nakagawa N, Mizuno M, Kato S, et al. Demographic, clinical characteristics and treatment outcomes of immune-complex membranoproliferative glomerulonephritis and C3 glomerulonephritis in Japan: A retrospective analysis of data from the Japan Renal Biopsy Registry. PLoS One. 2021;16(9):e0257397.

24. Ravindran A, Fervenza FC, Smith RJ, De Vriese AS, Sethi S. C3 glomerulopathy: ten years’ experience at Mayo Clinic. Elsevier; 2018:991–1008.

25. Chauvet S, Hauer JJ, Petitprez F, et al. Results from a nationwide retrospective cohort measure the impact of C3 and soluble C5b-9 levels on kidney outcomes in C3 glomerulopathy. Kidney international. 2022;102(4):904–916.

26. Caravaca-Fontán F, Díaz-Encarnación M, Cabello V, et al. Longitudinal change in proteinuria and kidney outcomes in C3 glomerulopathy. Nephrology Dialysis Transplantation. 2022;37(7):1270–1280.

27. Stevens LA, Greene T, Levey AS. Surrogate end points for clinical trials of kidney disease progression. Clinical Journal of the American Society of Nephrology. 2006;1(4):874–884.

28. Levey AS, Gansevoort RT, Coresh J, et al. Change in albuminuria and GFR as end points for clinical trials in early stages of CKD: a scientific workshop sponsored by the National Kidney Foundation in collaboration with the US Food and Drug Administration and European Medicines Agency. American journal of kidney diseases. 2020;75(1):84–104.

29. Nester C, Decker DA, Meier M, et al. DEVELOPING THERAPIES FOR C3G: REPORT OF THE KIDNEY HEALTH INITIATIVE C3G TRIAL ENDPOINTS WORK GROUP. Clinical Journal of the American Society of Nephrology. 2024:10.2215.

30. Grams ME, Sang Y, Ballew SH, et al. Evaluating glomerular filtration rate slope as a surrogate end point for ESKD in clinical trials: an individual participant meta-analysis of observational data. LWW; 2019. p. 1746–1755.

31. Inker LA, Heerspink HJ, Tighiouart H, et al. GFR slope as a surrogate end point for kidney disease progression in clinical trials: a meta-analysis of treatment effects of randomized controlled trials. LWW; 2019. p. 1735–1745.

32. Inker LA, Collier W, Greene T, et al. A meta-analysis of GFR slope as a surrogate endpoint for kidney failure. Nature medicine. 2023;29(7):1867–1876.

33. Heerspink HJ, Greene T, Tighiouart H, et al. Change in albuminuria as a surrogate endpoint for progression of kidney disease: a meta-analysis of treatment effects in randomised clinical trials. The lancet Diabetes & endocrinology. 2019;7(2):128–139.

34. Thompson A, Carroll K, Inker LA, et al. Proteinuria reduction as a surrogate end point in trials of IgA nephropathy. Clinical journal of the American Society of Nephrology: CJASN. 2019;14(3):469.

35. Inker LA, Mondal H, Greene T, et al. Early change in urine protein as a surrogate end point in studies of IgA nephropathy: an individual-patient meta-analysis. American Journal of Kidney Diseases. 2016;68(3):392–401.

36. Prentice RL. Surrogate endpoints in clinical trials: definition and operational criteria. Statistics in medicine. 1989;8(4):431–440.

37. Inker LA, Heerspink HJ, Tighiouart H, et al. Association of treatment effects on early change in urine protein and treatment effects on GFR slope in IgA nephropathy: an individual participant meta-analysis. American Journal of Kidney Diseases. 2021;78(3):340–349. e1.

38. Wong K, Pitcher D, Braddon F, et al. Description and Cross-Sectional Analyses of 25,880 Adults and Children in the UK National Registry of Rare Kidney Diseases Cohort. Kidney International Reports. 2024;

39. Levey AS, Stevens LA, Schmid CH, et al. A new equation to estimate glomerular filtration rate. Annals of internal medicine. 2009;150(9):604–612.

40. Schwartz GJ, Mun A, Schneider MF, et al. New equations to estimate GFR in children with CKD. Journal of the American Society of Nephrology. 2009;20(3):629–637.

41. Levin A, Agarwal R, Herrington WG, et al. International consensus definitions of clinical trial outcomes for kidney failure: 2020. Kidney international. 2020;98(4):849–859.

42. Wong K, Pitcher D, Braddon F, et al. Effects of rare kidney diseases on kidney failure: a longitudinal analysis of the UK National Registry of Rare Kidney Diseases (RaDaR) cohort. The Lancet. 2024;403(10433):1279–1289.

43. NHS. Blood and Transplant (2019) Annual Report on Kidney Transplantation: Report for 2018/19) Available from http://www.odt.nhs.uk/uk-transplant-registry/organ-specificreports/ [Accessed 3rd January 2024].

44. NHS. Blood and Transplant (2014) Annual Report on Kidney Transplantation: Report for 2013/14) Available from http://www.odt.nhs.uk/uk-transplant-registry/organ-specificreports/ [Accessed 3rd January 2024].

45. Khandelwal P, Bhardwaj S, Singh G, Sinha A, Hari P, Bagga A. Therapy and outcomes of C3 glomerulopathy and immune-complex membranoproliferative glomerulonephritis. Pediatric Nephrology. 2021;36:591–600.

46. FDA-NIH. Biomarker Working Group: BEST (Biomarkers, EndpointS, and other Tools) resource [Internet], Silver Spring, MD, Food and Drug Administration (US), 2016. Available at: http://www.ncbi.nlm.nih.gov/books/NBK326791/. Accessed June 24, 2024.

