## Supplemental Material for "Quantifying association of early proteinuria and eGFR changes with long-term kidney failure hazard in C3G and IC-MPGN"

Supplemental Table 1. Baseline characteristics and clinical outcomes of C3 Glomerulonephritis and Dense Deposit Disease

| | C3 Glomerulonephritis | | Dense Deposit Disease | |
|---|---|---|---|---|
| | N=99 | (%)* | N=54 | (%)* |
| **Age at diagnosis (years)** | | | | |
| Median (IQR) | 24 (12 - 46) | | 13 (8 – 20) | |
| Pediatric (<18 years) | 41 | 41 | 38 | (70) |
| **Sex** | | | | |
| Female | 43 | 43 | 23 | (43) |
| **Ethnicity** | | | | |
| White | 75 | 83 | 35 | (80) |
| **UPCR** (Median IQR, g/g) | | | | |
| Diagnosis | 3.8 (1.2 - 7.4) | | 3.4 (0.9 – 4.9) | |
| 6-month | 1.6 (0.8 – 3.7) | | 0.7 (0.1 – 3.0) | |
| 12-month | 2.0 (0.7 - 4.0) | | 0.9 (0.1 – 2.7) | |
| **eGFR at diagnosis** | | | | |
| Median (IQR), mL/min/1.73 $m^2$ | 70 (40 - 99) | | 87 (54- 107) | |
| **Serum albumin at diagnosis** | | | | |
| Median (IQR), g/dL | 3.2 (2.5 – 3.8) | | 2.6 (2.3 – 3.8) | |
| **Complement C3 levels at diagnosis** | | | | |
| Median (IQR), g/L | 0.29 (0.19 – 0.41) | | 0.18 (0.11 – 0.45) | |
| **Complement C4 levels at diagnosis** | | | | |
| Median (IQR), g/L | 0.18 (0.11 – 0.28) | | 0.19 (0.15 – 0.26) | |
| **Kidney Failure event** | | | | |
| Yes | 39 | (39) | 21 | 39 |
| **Time to kidney failure C3GN + DDD** | | | | |
| Median (IQR) (years) | 15.7 (10.2 – NE) | | | |
| **Immunosuppression within 1 year of diagnosis** | | | | |
| Yes | 26 | (36) | 21 | 47 |

*Percentages are proportions of those with data available. C3GN- C3 Glomerulonephritis, DDD- Dense Deposit Disease, IQR- interquartile range, NE- Not evaluable.

Supplemental Table 2. Urine protein creatinine ratio (UPCR) changes and hazard ratio of kidney failure definitive C3 and IC-MPGN subgroups

| IC-MPGN cohort | | 50% decline in UPCR and hazard ratio of KF | | | 0.44g/g (50mg/mmol) reduction in time averaged UPCR and hazard ratio of KF | | |
|---|---|---|---|---|---|---|---|
| **Timepoint from** | **Timepoint to** | **N** | **Unadjusted HR*** | **P value** | **N** | **Unadjusted HR*** | **P value** |
| Diagnosis | 6 months | 35 | 0.67 (0.49 - 0.91) | 0.01 | 35 | 0.88 (0.90 - 0.96) | 0.004 |
| Diagnosis | 1 year | 29 | 0.13 (0.04 - 0.47) | 0.002 | 29 | 0.51 (0.30 - 0.86) | 0.01 |
| 6 months | 1 Year | 23 | 0.46 (0.27 - 0.78) | 0.04 | 25 | 0.49 (0.28 - 0.88) | 0.02 |
| C3G (C3GN/DDD) cohort | | 50% decline in UPCR and hazard ratio of KF | | | 0.44g/g (50mg/mmol) reduction in TA- UPCR and hazard ratio of KF | | |
| **Timepoint from** | **Timepoint to** | **N** | **Unadjusted HR*** | **P value** | **N** | **Unadjusted HR*** | **P value** |
| Diagnosis | 6 months | 33 | 1.05 (0.47 - 2.32) | 0.92 | 36 | 0.97 (0.88 - 1.06) | 0.44 |
| Diagnosis | 1 year | 28 | 0.57 (0.31 - 1.02) | 0.06 | 29 | 0.85 (0.75 - 0.97) | 0.01 |
| 6 months | 1 Year | 30 | 0.90 (0.78 - 1.03) | 0.14 | 30 | 0.86 (0.76 - 0.99) | 0.03 |

* No adjustment possible due to small numbers IC-MPGN- Immune Complex Membranoproliferative Glomerulonephritis, KF – kidney failure, HR- Hazard ratio, C3G- C3 Glomerulopathy, C3GN- C3 Glomerulonephritis, DDD- Dense Deposit Disease

Supplemental Figure 1. Kaplan-Meier of time to 1st kidney transplant failure comparing C3G, IC-MPGN and full cohort

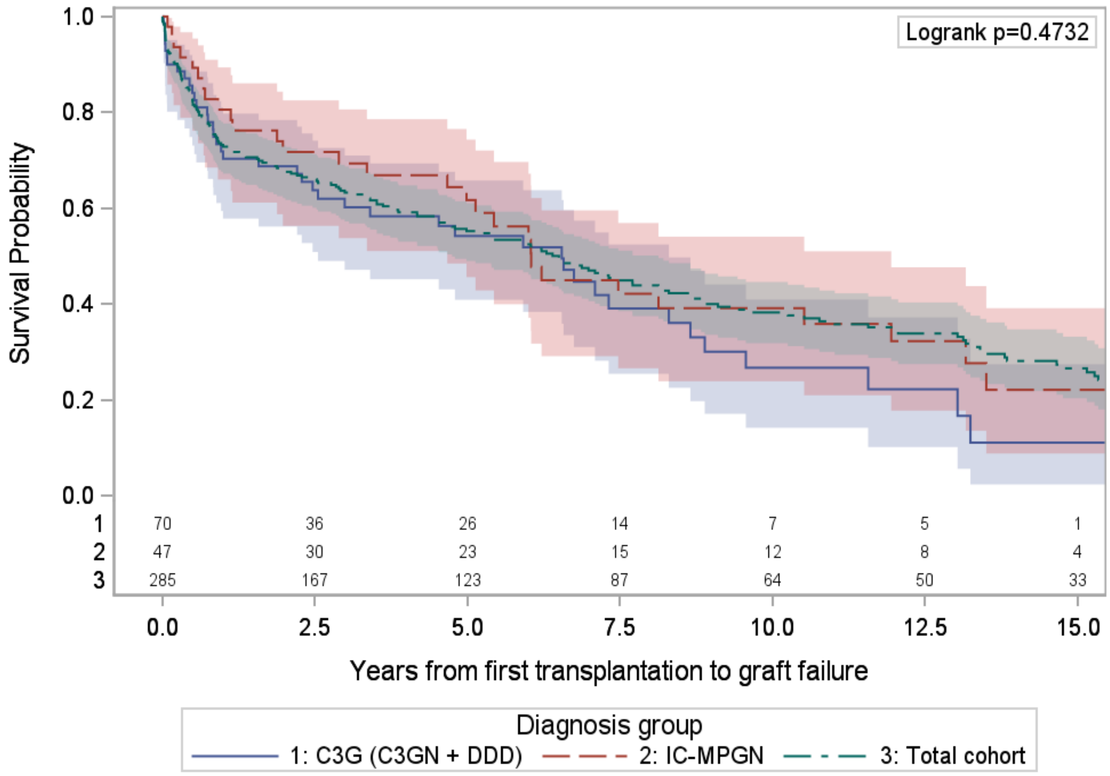

Supplemental Figure 2. Forest plot of eGFR slope within 2 years of diagnosis and risk of kidney failure (KF) for definitive C3G subgroup (Panel A) and definitive IC-MPGN subgroup (Panel B)

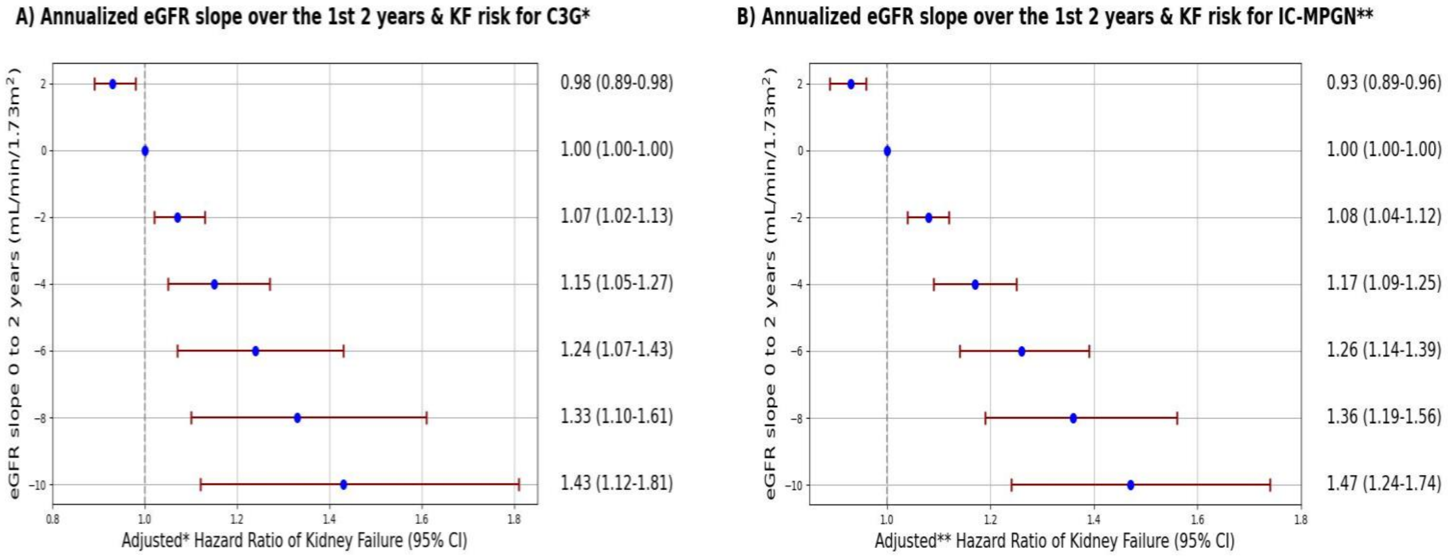


*Adjusted for age & baseline eGFR **Adjusted for sex & baseline eGFR

Supplemental Figure 3. Forest plot of 2-year eGFR slope for full cohort of prevalent patients (minimum 1 year from diagnosis) with an eGFR <60ml/min/1.73m$^2$ and risk of kidney failure

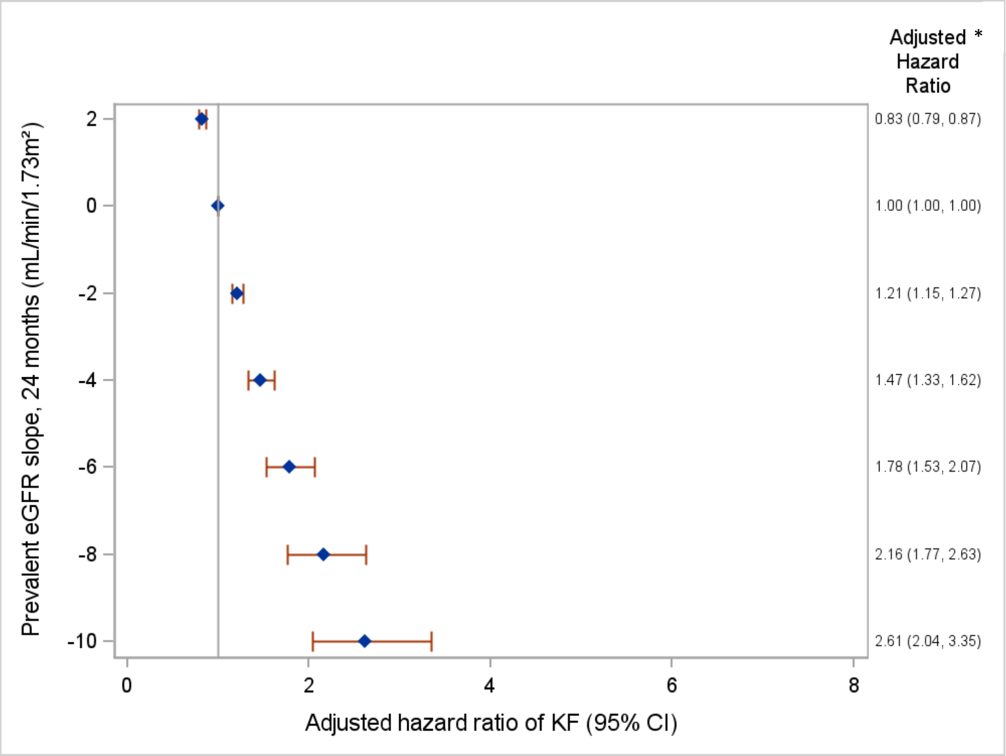


*Adjusted for baseline eGFR, sex and age

Supplemental Figure 4. Distribution of percentage change in eGFR between 0 and 2 years for full cohort

**Histogram of percentage change in eGFR between 0 and 2 years**

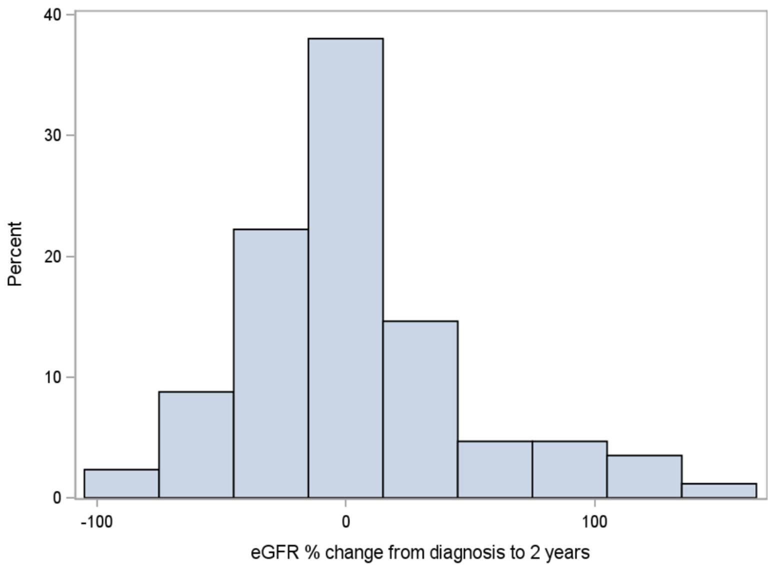

Supplemental Figure 5. Distribution of UPCR measurements for full cohort (Panel A) and on a log scale (Panel B)

Panel A: Distribution of UPCR measurements

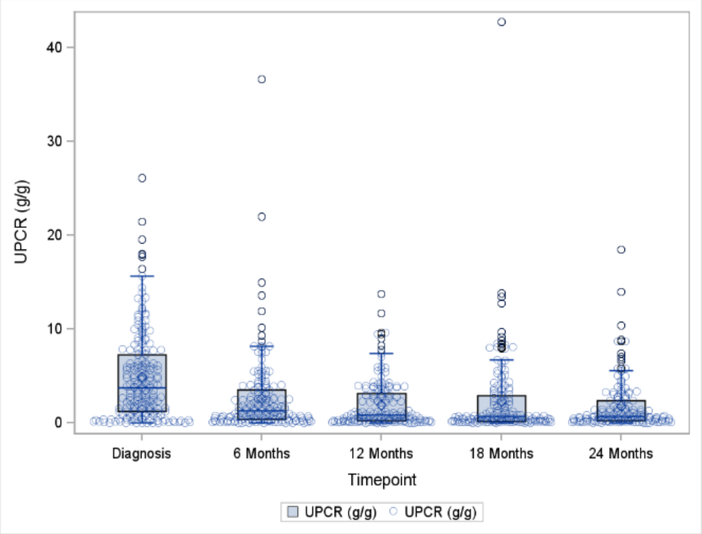


Panel B: Distribution of UPCR measurements on a Log scale

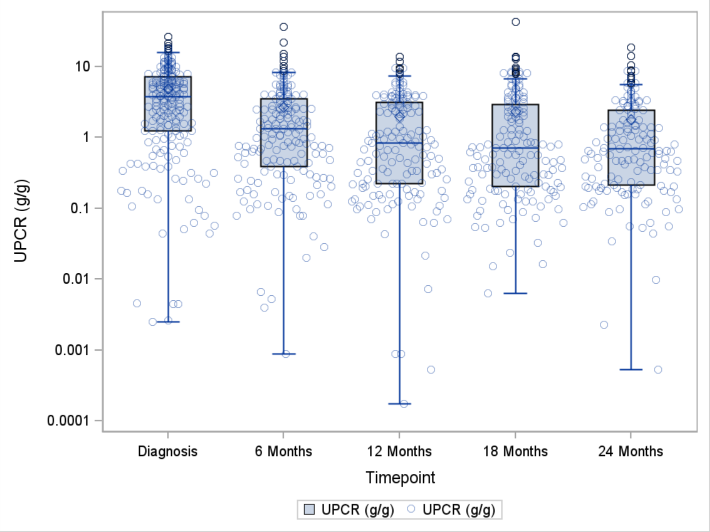

Supplemental Figure 6. Forest plot of percentage in UPCR 0 to 12 months and unadjusted risk of kidney failure (KF) for definitive C3G subgroup (Panel A) and definitive IC-MPGN subgroup (Panel B)

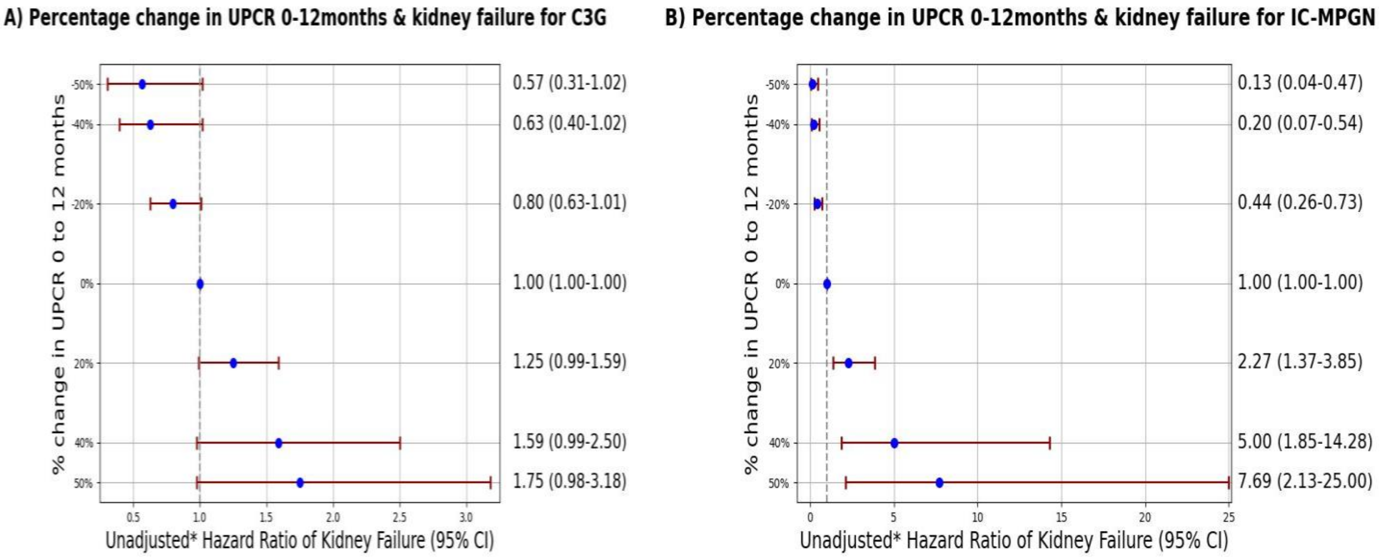


* No adjustment possible due to small numbers

Supplemental Figure 7. Distribution of percentage change in UPCR between 0 and 12 months for full cohort

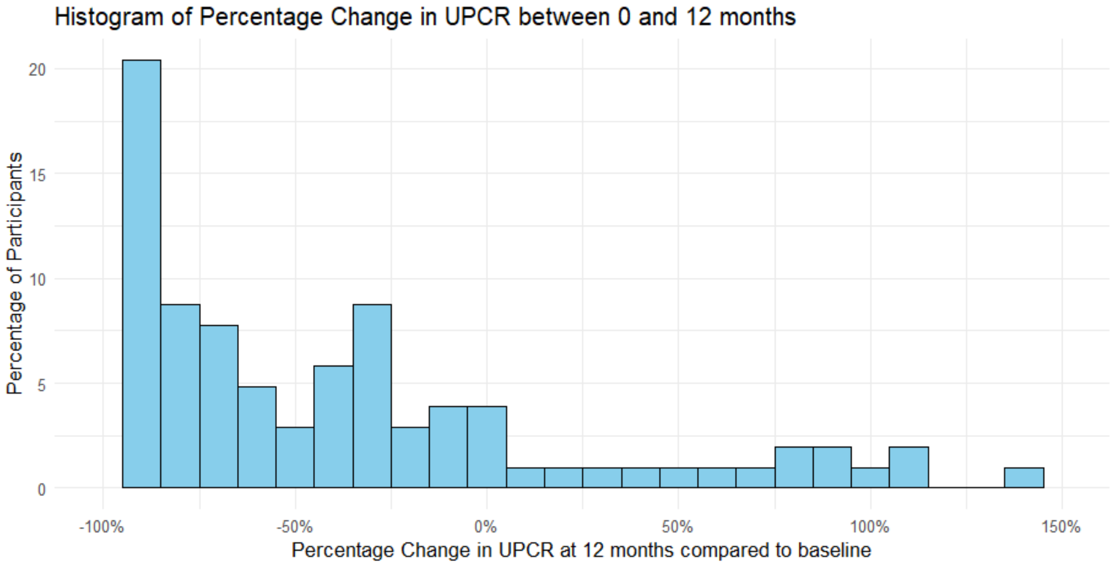
